# COVID-19 Vaccine Concerns about Safety, Effectiveness and Policies in the United States, Canada, Sweden, and Italy among Unvaccinated Individuals

**DOI:** 10.1101/2021.09.09.21263328

**Authors:** Rachael Piltch-Loeb, Nigel Harriman, Julia Healey, Marco Bonetti, Veronica Toffolutti, Marcia A Testa, Max Su, Elena Savoia

## Abstract

Despite the effectiveness of the COVID-19 vaccine, global vaccination distribution efforts have thus far had varying levels of success. Vaccine hesitancy remains a threat to vaccine uptake. This study has four objectives: 1) describe and compare vaccine hesitancy proportions by country; 2) categorize vaccine-related concerns; 3) rank vaccine-related concerns; and 4) compare vaccine-related concerns by country and hesitancy status in four countries- the United States, Canada, Sweden, and Italy. Using the Pollfish survey platform, we sampled 1000 respondents in Canada, Sweden, and Italy and 750 respondents in the United States between May 21-28, 2021. Results showed vaccine related concerns varied across three topical areas- vaccine safety and government control, vaccine effectiveness and population control, and freedom. For each thematic area, the top concern was statistically significantly different in each country and among the hesitant and non-hesitant subsamples within each county. Understanding the specific concerns among individuals when it comes to the COVID-19 vaccine can help to inform public communications and identify which, if any, salient narratives, are global.

## Introduction

The COVID-19 vaccine is the most effective prevention measure to reduce hospitalizations and mortality caused by SARS-CoV2 infections. As of September 2021, global vaccination distribution efforts have been underway for the past several months, with varying levels of success. In addition to the logistical hurdles related to the distribution of the vaccines to the public, vaccine hesitancy from certain segments of the population remains a challenge. Prior to the COVID-19 pandemic, vaccine hesitancy was on the rise, and in 2019 it was named by the WHO as one of the top ten threats to global health [1]. Despite variation in national approaches to vaccine promotion and vaccine policy, hesitancy is a concern for national and subnational governments challenged with developing and adapting communication strategies to a continuously evolving information environment. Globally, there have been several efforts to monitor trends in hesitancy for the COVID-19 vaccine. In a study conducted Rozek et al. in seventeen countries in May-June 2020 by (prior to vaccines being available), vaccine hesitancy rates ranged from 27% to 72% [2]. In a study conducted during a similar time frame across nineteen countries, Lazarus et al. found that overall 71% of respondents would be willing to take a vaccine, but that there was significant heterogeneity in vaccine hesitancy rates by country, demographic correlates of hesitancy, and policy perceptions [3]. In a more recent systematic review of global vaccine hesitancy rates, Sallam found relatively high levels of hesitancy (greater than 30%) in several European countries and the United States [4].

This study sought to characterize and compare the distribution of COVID-19 vaccine hesitancy across four countries: Canada, Italy, Sweden, and the USA, and identify and compare the top vaccine concerns among unvaccinated hesitant and non-hesitant individuals within each country. Each of the four countries shared similarities in their approach to procurement, distribution, and prioritization plans for the vaccines [5-8]. Previous research shows some of the top COVID-19 vaccine concerns in these countries were related to vaccine safety, speed of vaccine production, ingredients in the vaccine, adverse effects of the vaccine, political and financial objectives, and limited perceived risk of COVID-19 [9-11]. Though the reasons for vaccine hesitancy in each of these countries have been examined, prior research has not looked at the issue cross-nationally. To examine this issue, the current study had four objectives, namely: 1) describe and compare vaccine hesitancy proportions by country; 2) categorize vaccine-related concerns; 3) rank vaccine-related concerns; and 4) compare vaccine-related concerns by country and hesitancy status. Understanding the specific concerns among individuals when it comes to the COVID-19 vaccine can help to inform public communications and identify which, if any, salient narratives, are global.

## Methods

### Data collection

We conducted a cross-sectional, online survey via mobile devices on the Pollfish survey platform [12]. Pollfish pays mobile application developers to display the surveys within their applications. To incentivize participation, small monetary incentives are provided to randomly selected users who complete the surveys. The Pollfish platform uses random device engagement (RDE) to reach users engaged in using a mobile application who are identified only by a unique device ID. Pollfish has over 1 billion registered users worldwide. For this survey, a random sample of users who fit the study’s eligibility criteria was initially selected and data were collected between May 21-28, 2021. A sample of 1000 respondents was selected in Canada, Italy, and Sweden, and a sample of 750 respondents was selected in the USA. All samples had equally distributed quotas on gender and age groups (18-24, 25-34, 35-44, 45-54, and 55+). Individuals were eligible to participate if they were over 18 and had not yet been fully vaccinated. As the average time to thoughtfully complete the survey was estimated to take at the very least three minutes, we removed responses from all participants who took less than three minutes to complete the survey (n=87). After the removal of these respondents, the mean time to complete the survey was 9m55s (95% Prediction Interval: 3m00s to 29m00s). The study protocol and survey instrument were approved by the Harvard T.H. Chan School of Public Health Institutional Review Board. Participants reviewed information on the study before consenting to participate by clicking through to the next page and beginning the survey. No minors were involved in this study.

### Vaccine hesitancy

To define an individual as being vaccine-hesitant, we used the answer to the question “If you were offered a COVID-19 vaccine - at no cost to you - how likely are you to take it?” We set a dummy variable equal to 1 (“hesitant”) for those who answered, “I would not take it at the moment but would consider it later on,” or “Very unlikely,” or “Somewhat unlikely,” or “I am not sure,” or “Somewhat likely”. The variable was set equal to 0 for the individuals who picked the remaining possible answer, “Very likely” (“non-hesitant”).

### COVID-19 Vaccine Concerns

Respondents were asked to rate how much they agreed with a statement from 1 (strongly disagree) to 7 (strongly agree) of 19 potential concerns related to vaccine safety, effectiveness, distribution, and policy. Some of the items were designed to detect perception of misinformation (i.e., the vaccine containing a microchip to track the population). Each item was transformed from a 1-7 scale into a 1-10 scale for ease of presentation with 1 representing “low concern” and 10 “high concern”. See the Supplementary Material for the coding of the survey items. There were a total of 19 items, representing three vaccine hesitancy domains corresponding to 1) vaccine safety and government control; 2) vaccine effectiveness and population control; and 3) freedom based upon our initial conceptualization of the potential correlates of vaccine hesitancy.

### Statistical Analysis

To describe whether vaccine hesitancy varied by country we applied Kruskal-Wallis tests (objective 1). To categorize the vaccine concerns, we conducted a factor analysis to identify the underlying constructs and thematic grouping of survey items (objective 2). We then ranked these concerns by calculating the lowest mean agreement item within each thematic group based on the results of the factor analysis. We implemented repeated measures mixed models within these three groups to check that the means for each group of items differed in a statistically significant fashion. Repeated measures mixed models were used to test between-group (by country and hesitancy status) differences. (objective 3). We then conducted Kruskal-Wallis tests to describe whether the medians of the vaccine concerns varied by country for three groups of respondents: 1) overall sample, 2) vaccine-hesitant individuals, and 3) non-hesitant individuals. Finally, we describe the top vaccine concerns by the above grouping (objective 4).

## Results

### Objective 1 – Describe and Compare Vaccine Hesitancy Proportions by Country

Vaccine hesitancy by country is reported in Table 1. The median scores of vaccine hesitancy varied by country (Kruskal-Wallis p<0.001). The US had the highest percentage of vaccine-hesitant respondents (63%), followed by Sweden (49%), Italy (43%), and Canada (42%).

**Table 1.** Distribution of Vaccine Hesitancy by Country.

|  | <u>US</u> |  | <u>Canada</u> |  | <u>Sweden</u> |  | <u>Italy</u> |  | <u>Total</u> |  |
| --- | --- | --- | --- | --- | --- | --- | --- | --- | --- | --- |
|  | *N* | % | *N* | % | *N* | % | *N* | % | *N* | % |
| *I would not take it at the moment but would consider it later on* | 81 | 11% | 46 | 5% | 37 | 4% | 37 | 4% | 201 | 5% |
| *Very unlikely* | 133 | 18% | 83 | 8% | 62 | 6% | 37 | 4% | 315 | 9% |
| *Somewhat unlikely* | 41 | 6% | 31 | 3% | 44 | 5% | 30 | 3% | 146 | 4% |
| *I am not sure* | 86 | 12% | 113 | 11% | 152 | 16% | 100 | 10% | 451 | 12% |
| *Somewhat likely* | 118 | 16% | 140 | 14% | 176 | 18% | 222 | 23% | 656 | 18% |
| *Very likely* | 268 | 37% | 572 | 58% | 494 | 51% | 560 | 57% | 1,894 | 52% |
| *Total* | 727 |  | 985 |  | 965 |  | 986 |  | 3,663 |  |

### Objective 2 – Categorize Vaccine-Related Concerns

#### Underlying Constructs and Grouping of Survey Items

Initially, three factor analyses were performed on each of the originally hypothesized domains of vaccine concern items to confirm the unidimensionality of each set of questions as presented and grouped in the survey. Results supported the unidimensionality of two of the three domains while the third domain identified two factors (See supplemental material for the statistical output). To explore the measurement structure of the two-factor domain, an exploratory factor analysis, using all nineteen items was performed. KMO measure of sampling adequacy (0.94) and Bartlett’s test for sphericity (p<0.01) indicated that the nineteen items were suitable for factor analysis. Results identified three factors with eigenvalues greater than one (53.17% variance explained) after oblique Promax rotation (Promax power=1). These factors provided the basis for grouping the nineteen questions in the questionnaire into three updated domains which after examining the item content with the domains, we named: 1) vaccine effectiveness & population control, 2) vaccine safety & government control, and 3) freedom. (See Supplemental material for the output of the exploratory factor analysis).

### Objective 3 – Rank Vaccine-Related Concerns

The top overall concern for each of the three domains of vaccine concerns is displayed in Table 2. Regarding the group of items describing concerns about “vaccine effectiveness and population control”, the top concern was that “an elite group would achieve financial power through the vaccine” (mean=6.1, sd=2.7). For the group of items describing concerns related to “vaccine safety and government control”, across the entire sample, the top concern was that “the vaccine could cause other diseases” (mean agreement=5.3, sd=2.6). Finally, respondents’ top concern among items describing freedom was that “people should be free to decide if they get vaccinated or not with no consequences for their job or personal life “(mean=6.3, sd=2.8). With the exception of the vaccine safety and government control questions among non-hesitant US respondents, repeated measures models indicated that the means for each set of concerns (vaccine safety & government control, vaccine effectiveness & population control, and freedom) were significantly different by country and hesitancy status.

**Table 2.** Top Overall Concerns.

| *Topic* | *Reason* | *Mean (SD)* |
| --- | --- | --- |
| *Vaccine Safety and Government Control* | *The vaccine can cause other diseases* | 5.3 (2.6) |
| *Vaccine Effectiveness and Population Control* | *There is no elite group that will achieve financial power if people are getting vaccinated* | 6.1 (2.7) |
| *Freedom* | *People should be free to decide if they get vaccinated or not with no consequences for their job or personal life* | 6.3 (2.8) |

### Objective 4 – Vaccine Concerns by Country and Hesitancy Status

#### Distribution of Vaccine Concerns across Countries

Across the entire sample, there were different medians for each question (Kruskal-Wallis p<0.01) by country. Compared to the other countries, the United States had the lowest mean value for 14 of the 19 questions. Among those who were not vaccine-hesitant, the only concern that had the same median in each country was the level of concern about the vaccine being used by governments to limit civil rights (Kruskal-Wallis p>0.05). Across the “hesitant” subsample, there was the same median level of concern in each country related to if the vaccine can cause other diseases, getting COVID-19 rather than the vaccine, and if an elite group will achieve financial power if people are getting vaccinated (Kruskal-Wallis p>0.05).

#### Top Vaccine Concerns by Country

The top vaccine concerns by country and hesitancy status are reported in Figures 1-3. Below we highlight the key differences across countries and hesitancy status.

**Figure 1.**
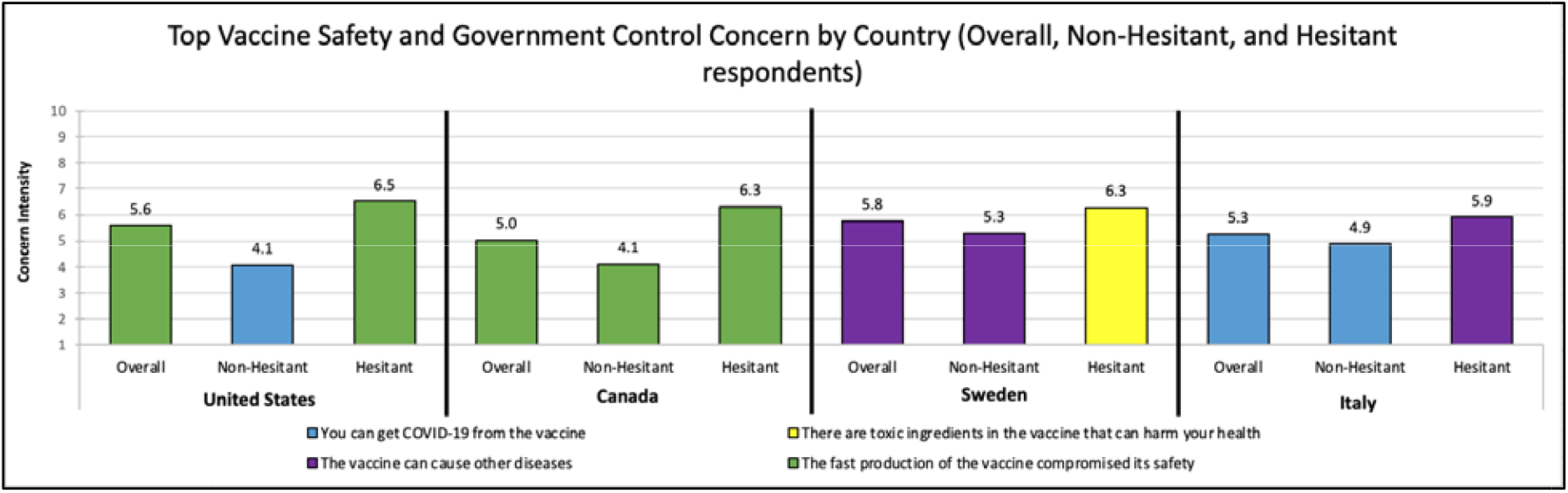
Top Vaccine Safety and Government Control Concern by Country and Hesitancy Status.

**Figure 2.**
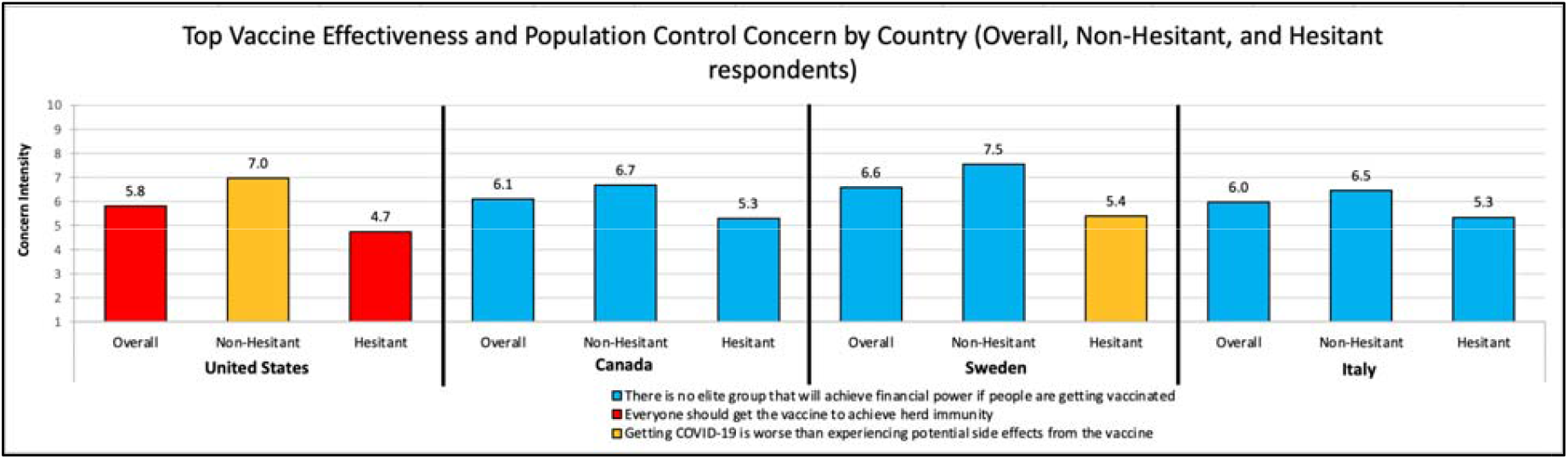
Top Vaccine Effectiveness and Population Control Concern by Country and Hesitancy Status.

**Figure 3.**
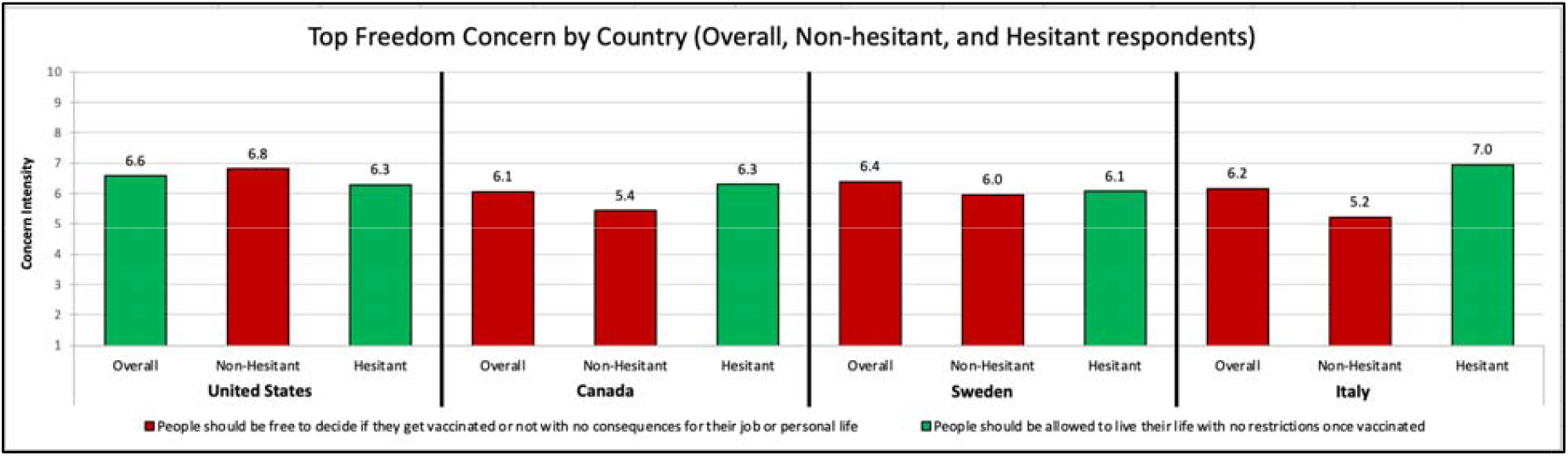
Top Freedom Concern by Country and Hesitancy Status.

#### Vaccine Safety and Government Control

Figure 1 describes the top vaccine safety and government control concerns by country. Respondents from the US and Canada shared the same top vaccine safety and government control concern: *the fast production of the vaccine compromised its safety*. Respondents from Sweden (*the vaccine can cause other diseases)* and Italy *(you can get COVID-19 from the vaccine*) were both unique in their top vaccine safety and government control concerns. Among respondents who were not vaccine-hesitant, the US and Italy shared the same top concern (*you can get COVID-19 from the vaccine*), while Canada (*the fast production of the vaccine compromised its safety) and Sweden (the vaccine can cause other diseases*) were both unique. Among the hesitant, the US and Canada shared the same top concern (*the fast production of the vaccine compromised its safety*), while Sweden (*there are toxic ingredients in the vaccine*) and Italy (*the vaccine can cause other diseases*) were again unique.

#### Vaccine Effectiveness and Population Control

Figure 2 describes the top vaccine effectiveness and population control concerns by country. Respondents from Canada, Sweden, and Italy shared the same top vaccine effectiveness and population control concern (*there is no elite group that will achieve financial power if people are getting vaccinated*). The US (*everyone should get vaccinated to achieve herd immunity*) was unique. Among those who were not vaccine-hesitant, respondents from Canada, Sweden, and Italy again shared the same top concern (*there is no elite group that will achieve financial power if people are getting vaccinated*), while the US was unique (*getting COVID-19 is worse than the potential side effects of the vaccine*). Among the hesitant, Canada and Italy shared the same top concern (*there is no elite group that will achieve financial power if people are getting vaccinated*), while the US (*everyone should get vaccinated to achieve herd immunity*) and Sweden (*getting COVID-19 is worse than the potential side effects of the vaccine*) were both unique.

#### Freedom

Figure 3 describes the top freedom concerns by country. Of the four freedom questions, only two were in the top concerns of any country or hesitancy-status group: 1) people should be free to decide if they get vaccinated or not with no consequences for their job or personal life and 2) people should be allowed to live their life with no restrictions once vaccinated. Respondents from Canada, Sweden, and Italy shared the same top freedom concern (*people should be free to decide if they get vaccinated or not*), while the US was unique (*people should be allowed to live their life with no restrictions once vaccinated*). Among those who were not vaccine-hesitant, respondents from all four countries shared the same top freedom concern (*people should be free to decide if they get vaccinated or not*). Among the hesitant, all four countries again shared the same top freedom concern (*people should be allowed to live their life with no restrictions once vaccinated*).

## Discussion

The first key finding from our analysis is that concerns about the safety and effectiveness of the vaccine overlapped with concerns about population and government control as indicated by the results of the factor analysis. In addition, concerns derived from personal risk-benefit considerations (i.e., side-effects from the vaccine compared to effects of COVID-19) overlapped with concerns derived from perceptions of mis-disinformation (i.e., the government is using the vaccine for control). These findings underscore the complexity of the vaccine hesitancy construct which includes not only concerns about the vaccine as a pharmaceutical product and individual risk-benefit considerations but also concerns about the policies for its distribution. Some of these concerns have been the focus of mainstream media reports, such as “is herd immunity achievable”. Other concerns have been fueled by alternative media sources such as “the vaccine includes a microchip”. There is complexity of individuals’ exposure to an information environment where misinformation is covering topics ranging from effectiveness, safety to policies.

There were distinct findings across each of the themes we identified. On the first factor, related to safety and government control, there was an apparent distinction between respondent’s concerns in the European countries (Italy and Sweden) and the North American countries (the US and Canada). In relation to safety and government control, the top concerns in the European countries were focused on health issues that may arise from the vaccine-either because of concerns that the vaccine could include toxic ingredients or concerns that the vaccine could cause other health issues. This is consistent with previous findings from a European survey on COVID-19 vaccine hesitancy. Vaccine ingredients and potential adverse side effects from the vaccine were found as top concerns for Italian respondents [11]. Additionally, prior work with a sample of Swedish respondents found there was the perception that safety considerations were bypassed during COVID-19 vaccine production [11]. In contrast, the top concern in the North American countries focused on the fast production of the vaccine. This has been identified by other authors as a reason for vaccine hesitancy in both the US and Canada. Researchers in Canada found that the top concern about the COVID-19 vaccine was related to safety due to the speed of production, as well as about possible side effects from vaccination [9]. In the USA, prior research showed one of the top concerns among unvaccinated people was also related to COVID-19 vaccine safety and possible adverse side effects [10]. This distinction between the European and the North American countries may be related to a variety of factors that require further investigation. One hypothesis is that the media coverage and dominant narratives in North American coverage, were different from those in Europe, resulting in distinct dominant narratives. Future analyses will explore how differences in the sample composition, political leanings, and media consumption may relate to particular beliefs.

The key finding on the second factor was that the US respondents were an outlier in the level of concern about vaccine effectiveness and population control. Respondents overall and among the hesitant did not agree that vaccination would achieve herd immunity. This may be a reflection of the difficulty in achieving herd immunity and lack of data on what level of coverage is needed or if it is even possible to achieve such a goal with an ever-mutating virus. In contrast, in Canada, Sweden, and Italy there was consistency that the greatest concern had to do with elites benefitting from the vaccine rollout. In Europe, especially in relation to the AstraZeneca vaccine, news reports highlighted political elites extolling power to procure and distribute vaccines [13-15]. This may have contributed to the concern overall. In Canada, there appeared to have been a recognition of this potential concern, so much so that politicians were not prioritized for vaccination during the rollout, and political elites from multiple parties directed their early support behind vaccination [16].

Finally, regarding freedom concerns, we found the greatest consistency across countries and hesitancy groups. Across all four countries, there were the same top two concerns-that there should be freedom of choice to be vaccinated and freedom of movement when vaccinated. Among the hesitant, the top concern across all four nations related to freedom of movement once vaccinated. The sentiment being that vaccination status should not be tied to movement. This similarity overall and among the hesitant suggests there may be salient global narratives that know no borders and reflect the spread of ideas online or in media. This is also a lesson for policymakers, that regardless of nation of origin, consistent concerns remain among the unvaccinated.

This analysis is limited in that it is a descriptive snapshot of hesitancy concerns. Since this study was conducted, the vaccine rollout has expanded especially in Canada and in Europe. However, as of the end of Summer 2021, there remains nearly thirty percent of the population who are unvaccinated, many of whom do not intend to get the vaccine [17]. Unvaccinated individuals have varied concerns, but it is in the interest of risk communicators and policy makers to focus on the concerns of the hesitant. This study highlights the distinct concerns among that group. Risk communication must be clear and concise to be effective. The results can be used by policy makers in each country who are aiming to craft the key message to deliver to their population of interest. For policy makers within one of the countries where this study was conducted, we have demonstrated key messages to highlight, and for international organizations, our results also show there are global themes-especially when it comes to personal freedom-that should be incorporated into risk communication.

## Data Availability

Data is available upon request from the authors.

## Acknowledgement

This project received funding from the NATO Science for Peace and Security Programme (grant number SPS.MYPG5556), the US Department of Homeland Security (DHS), Science and Technology Directorate (Cooperative Agreement Number: 2015-ST-108-FRG005) and Swedish Contingency Agency (MSB). The content of this manuscript as well as the views and discussions expressed are solely those of the authors and do not necessarily represent the official views of any of the above institutions, nor does the mention of trade names, commercial practices, or organizations imply endorsement by the US government.

## Supplementary Material – Confirmatory Factor Analysis

| Item | Vaccine Safety and Government Control |
| --- | --- |
| You can get COVID-19 from the vaccine itself (R) | 0.6668 |
| There are toxic ingredients in the vaccine that can harm your health (R) | 0.7663 |
| The vaccine can mess up your DNA (R) | 0.7268 |
| The vaccine can cause infertility (R) | 0.7359 |
| The vaccine can cause other diseases (R) | 0.7356 |
| The fast production of the vaccine compromised its safety (R) | 0.6907 |
| The vaccine is going to be used by Governments as a tool to limit our civil rights (right of assembly, right of movement, right of religion, etc.) | 0.6808 |

| Item | Vaccine Effectiveness and Population Control |
| --- | --- |
| The vaccine will work in protecting me from getting COVID-19 | 0.7406 |
| By taking the vaccine I will protect my friends and family from getting COVID-19 | 0.7926 |
| Everyone should get the vaccine to achieve herd immunity | 0.8098 |
| Getting COVID-19 is worse than experiencing potential side effects from the vaccine | 0.6863 |
| Natural remedies will not protect me from COVID-19 | 0.6385 |

| Item | Freedom - Factor 1 | Freedom - Factor 2 |
| --- | --- | --- |
| People should be free to decide if they get vaccinated or not with no consequences for their job or personal life | 0.807 |  |
| People should have the option to choose the vaccine brand they want | 0.7607 |  |
| People should be allowed to live their life with no restrictions once vaccinated | 0.437 |  |
| Healthcare professionals and scientists with concerns about the vaccine should have opportunities to share their opinions with the public | 0.6284 |  |
| There is no elite group that will achieve financial power if people are getting vaccinated |  | 0.6319 |
| There is no microchip with tracking capabilities inserted in the vaccine |  | 0.7415 |
| Everybody should have equal access to the most effective and safe vaccine regardless of income, race, or immigration status |  | 0.8364 |

## Supplementary Material – Exploratory Factor Analysis

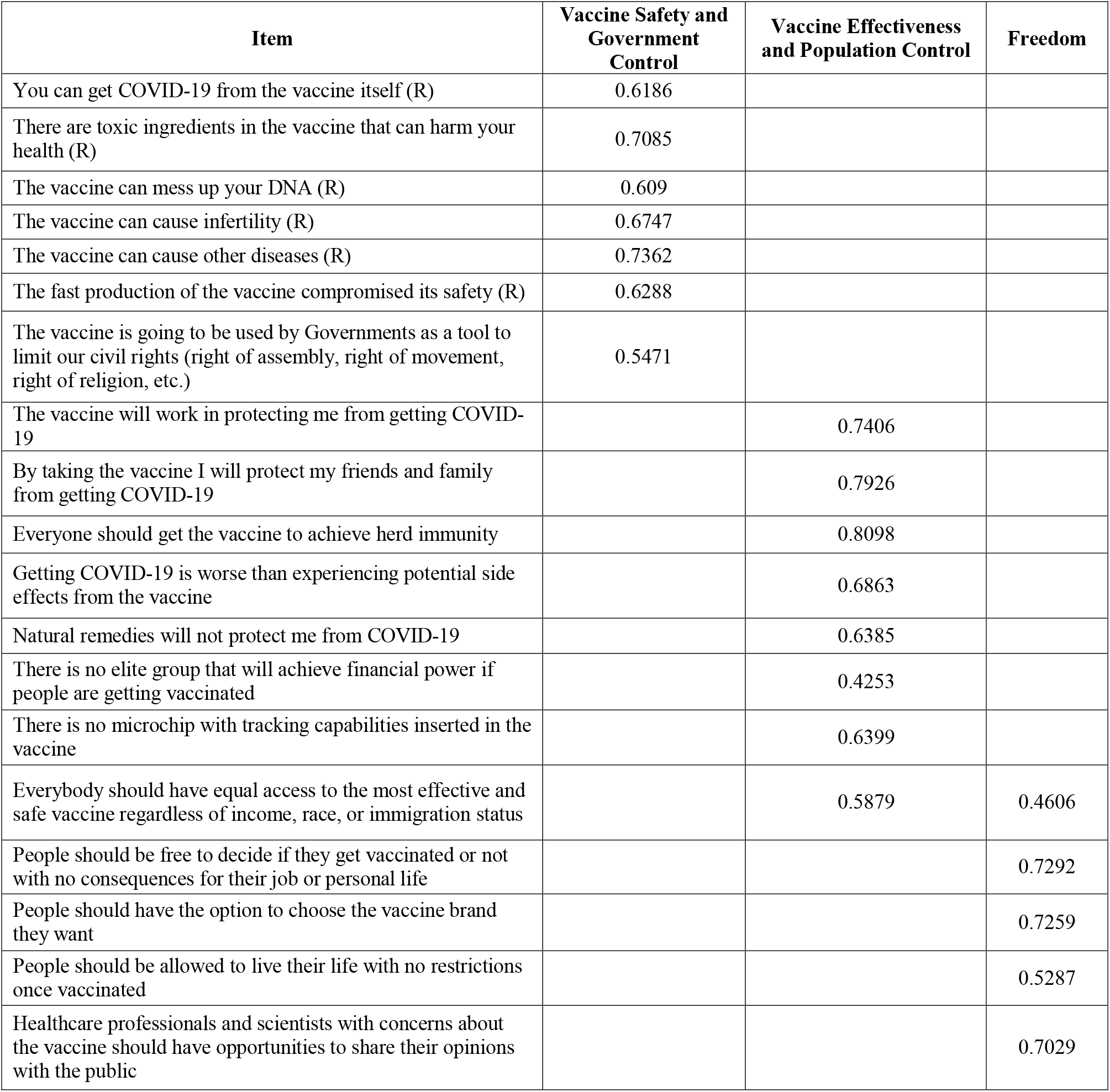
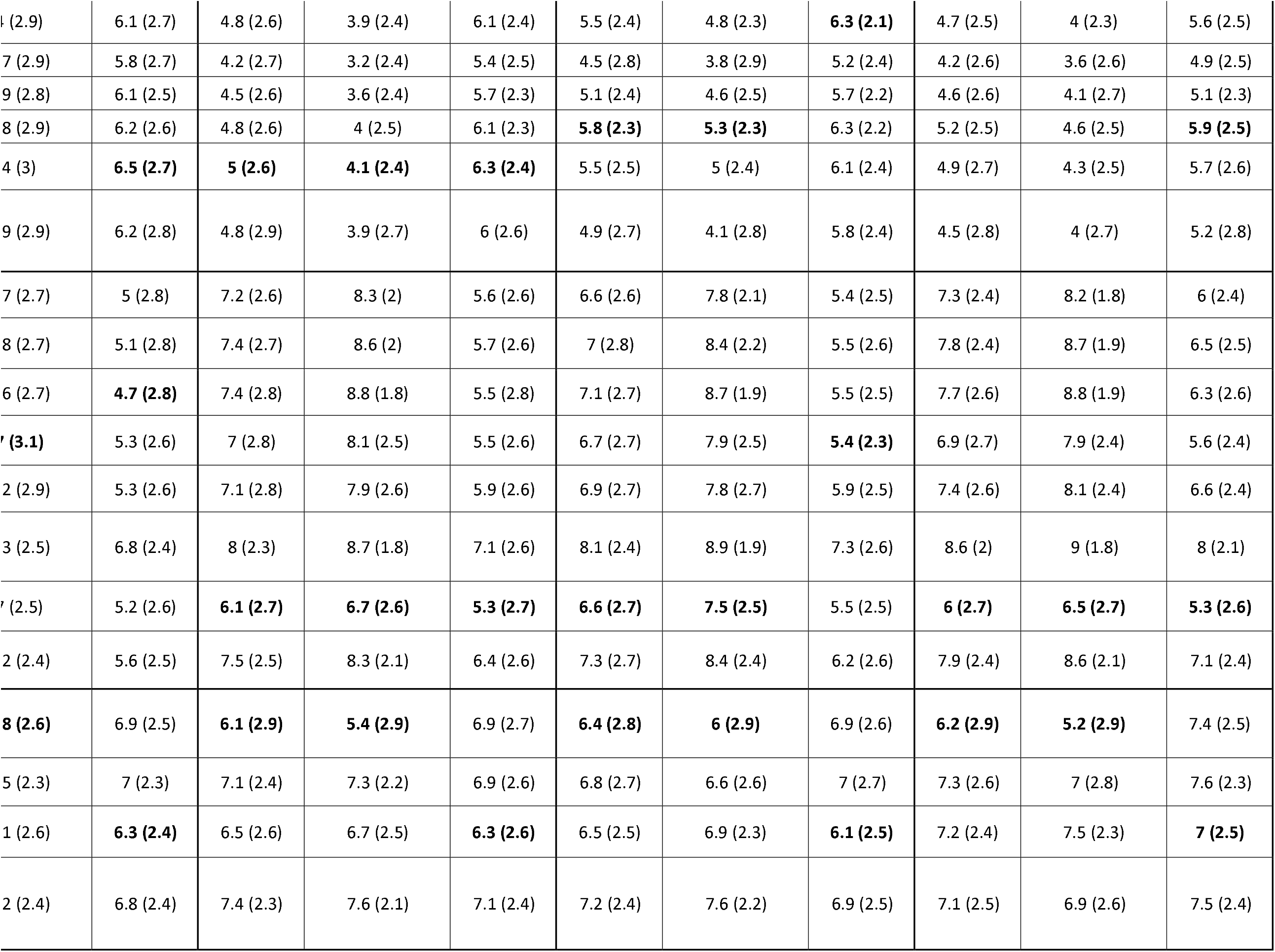

## Supplementary Material – Survey Items

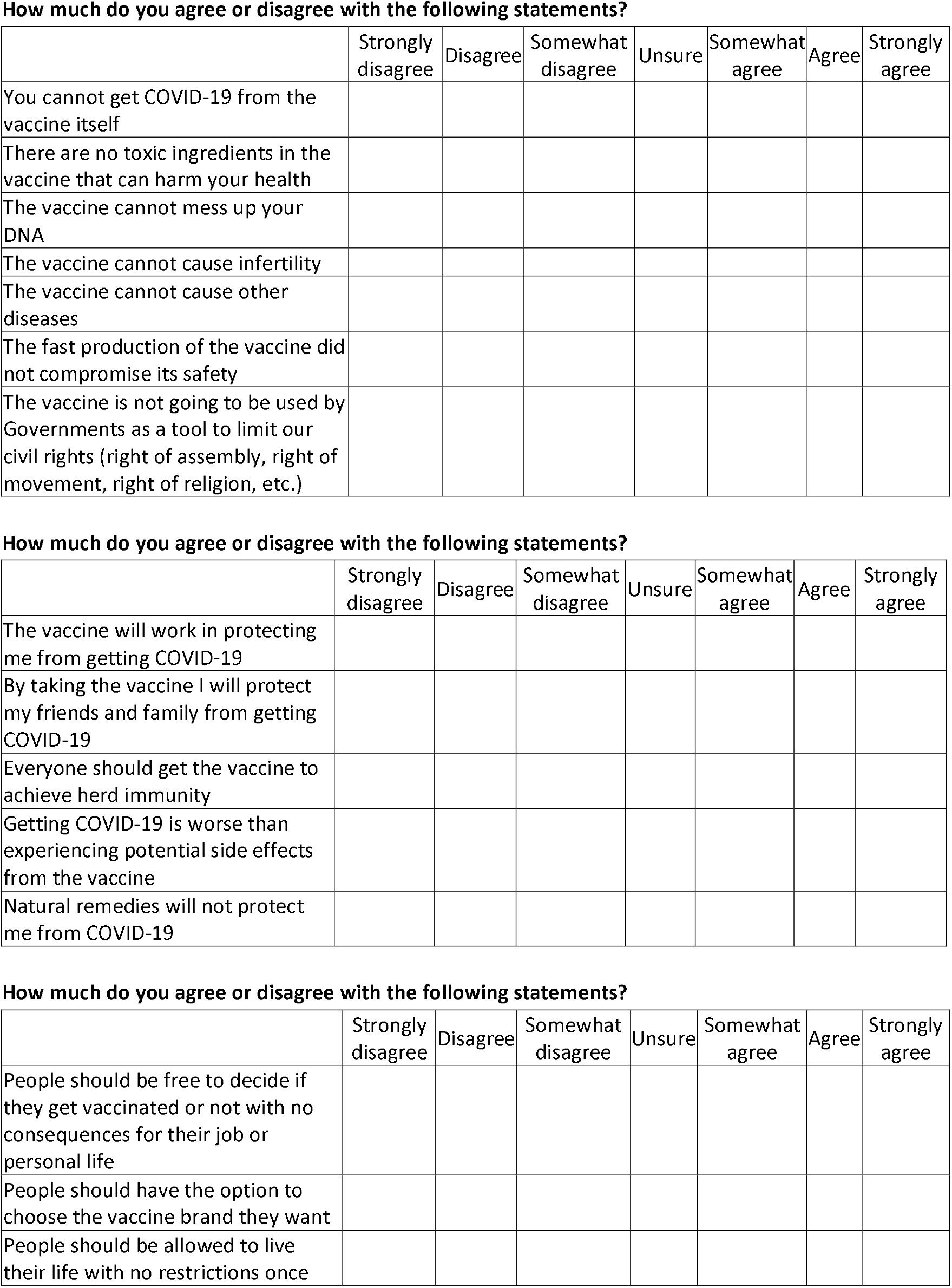
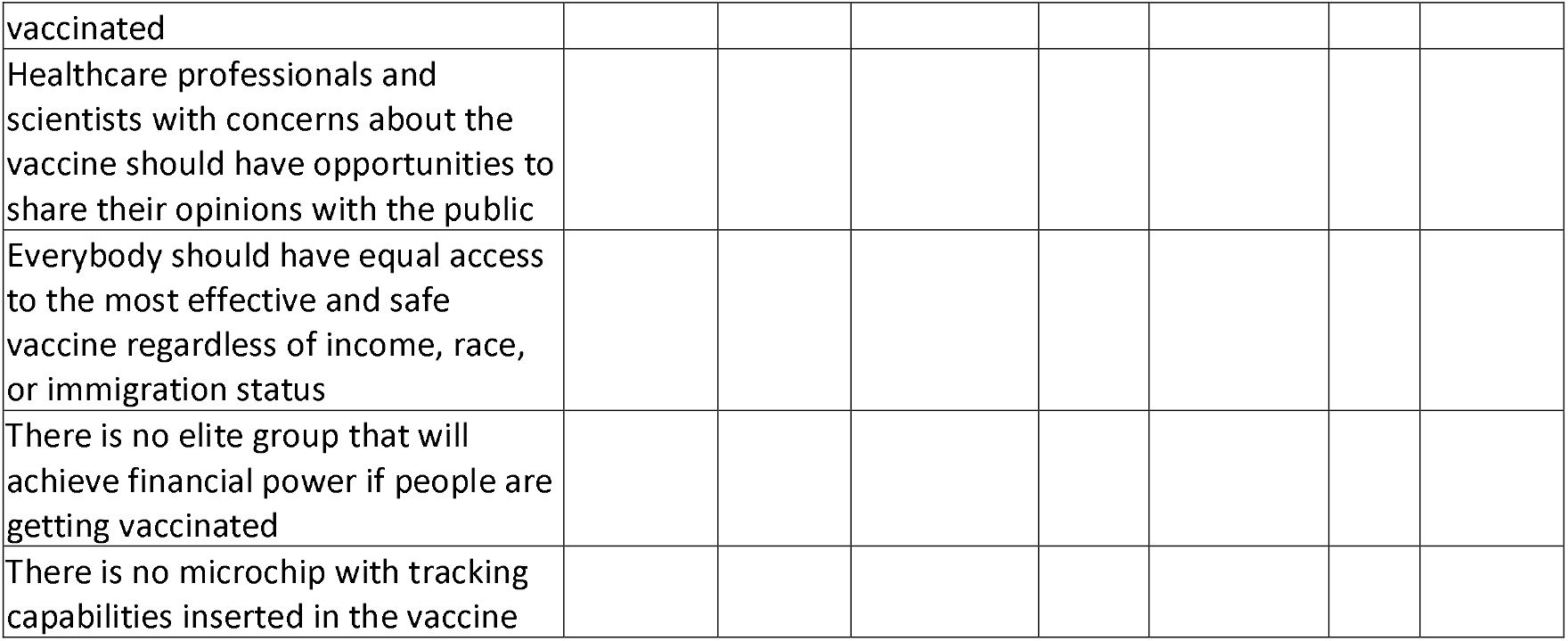

